# Strain-specific anti-RBD IgG antibody titers against the WT, XBB.1.5, JN.1, and KP.3 strains consistently correlate with neutralizing activity following SARS-CoV-2 XBB.1.5-adapted mRNA vaccination

**DOI:** 10.1101/2025.03.13.25323892

**Authors:** Takeyuki Goto, Yong Chong, Tomonori Sato, Naoki Tani, Shouta Saiki, Satoru Ishida, Naoki Kawai, Takuma Bando, Hideyuki Ikematsu

**Affiliations:** Medicine and Biosystemic Science, Kyushu University Graduate School of Medical Sciences, Fukuoka, Japan; Department of Clinical Immunology, Rheumatology, and Infectious Disease, Kyushu University Hospital, Fukuoka, Japan; Department of Infectious Diseases, Fukuoka City Hospital, Fukuoka, Japan; Vaccine R&D Laboratory, Shionogi & Co., Ltd., Osaka, Japan; Japan Physicians Association, Tokyo, Japan; Kawai Clinic, Gifu, Japan; Bando Clinic, Ishikawa, Japan; Ricerca Clinica Co., Fukuoka, Japan

**Keywords:** SARS-CoV-2, Receptor binding domain, IgG antibody titer, Neutralizing activity, Humoral immunity

## Abstract

**Introduction:** Data on the correlation between SARS-CoV-2 neutralizing activity and strain-specific anti-RBD IgG antibody (anti-RBD) titers is limited, particularly in the context of XBB.1.5-adapted vaccination.

**Methods:** A direct comparison of neutralizing activity, measured as 50% neutralization (NT_50_), and anti-RBD titers, measured using an ELISA, was conducted using serum samples collected in Japan before and after XBB.1.5-adapted mRNA vaccination.

**Results:** A total of 108 serum samples from 54 patients were analyzed. A strong correlation between neutralizing activity and anti-RBD titers was observed for the wild-type (WT), XBB.1.5, JN.1, and KP.3 strains (r = 0.94, 0.87, 0.86, and 0.82, respectively). This correlation persisted when stratifying pre- and post-vaccination samples (r = 0.92, 0.83, 0.85, and 0.82, respectively, for pre-vaccination samples and r = 0.96, 0.85, 0.82, and 0.75, respectively, for post-vaccination samples). Both NT_50_ and anti-RBD titers significantly increased against all four tested strains after vaccination (*p* < 0.001), with the highest fold change observed for the XBB.1.5 variant. Additionally, variant specificity, defined as the ratio of variant to WT values, significantly increased for XBB.1.5, JN.1, and KP.3 after vaccination in NT_50_ and was also observed in anti-RBD titers.

**Conclusions:** These findings, demonstrating a strong correlation with neutralizing activity not only against the WT strain but also against the XBB.1.5, JN.1, and KP.3 variants, suggest that strain-specific anti-RBD IgG antibody titers would be useful as an indicator of humoral immunity following XBB.1.5-adapted vaccination.

## Introduction

The evaluation of humoral immunity against SARS-CoV-2 has become increasingly complex due to the emergence of novel variants and the heterogeneous vaccination and infection histories of individuals. Neutralizing activity is widely recognized as a direct indicator of host humoral immunity against SARS-CoV-2 [1,2]. Measurement of anti-Spike IgG antibody (anti-S) or anti-RBD IgG antibody (anti-RBD) titers with an enzyme-linked immunosorbent assay (ELISA) provides a less labor-intensive and more cost-effective alternative [3]. Several studies have demonstrated the utility of such measurements in estimating neutralizing activity for the wild-type (WT) and Delta (B.1.617.2) strains [4–6]. Our previous study, conducted during the Omicron BA.1 (B.1.1.529) wave in Japan, demonstrated that strain-specific anti-S and anti-RBD IgG titers varied in a pattern similar to that of neutralizing activity among patients with different immune exposure backgrounds. Furthermore, integrating these titer measurements with neutralizing activity analysis offered additional qualitative insights [7].

Recently, the XBB.1.5-adapted vaccine has been widely administered in Japan, and the rollout of the JN.1-adapted vaccine is currently underway. Additionally, the KP.3 variant and its descendant lineages are spreading nationwide [8]. These Omicron subvariants reportedly harbor antigenic and structural features that facilitate evasion from host immunity targeting ancestral strains [9–13]. Despite this evolving landscape, neutralizing activity remains a key indicator of humoral immunity [14,15]. However, mainly because direct comparisons with neutralizing activity are limited, the alternative role of strain-specific anti-RBD titers for these emerging variants are yet to be fully confirmed.

To address this gap, we conducted a multicenter prospective study to directly compare neutralizing activity and strain-specific anti-RBD titers by analyzing serum samples collected before and after administration of the XBB.1.5-adapted vaccine in a clinical setting. Our objective was to assess the correlation between strain-specific neutralizing activity and anti-RBD titers for both the original and emerging strains and to add insights into the humoral immune response to these variants following XBB.1.5-adapted vaccination.

## Methods

### Participants

Participant enrollment occurred between September 2023 and March 2024 at four participating clinics. Serum samples were collected from participants who visited an enrolled clinic before and after receiving the XBB.1.5-adapted SARS-CoV-2 mRNA vaccination. Informed consent was obtained from participants in a preceding study that assessed symptoms after vaccination. In this study, opt-out was conducted for additional serological analysis. The attending physician reviewed past vaccination and infection history for SARS-CoV-2. Participants who contracted SARS-CoV-2 between the pre- and post-vaccination sampling points were excluded. Both the previous and this study were approved by the institutional review board of Hara-Doi Hospital (Approval Dates: September 19, 2023, and August 20, 2024, respectively).

### Measurement of neutralizing activity

SARS-CoV-2 pseudo virus generation and neutralization assays were conducted as described previously [16]. Briefly, lentivirus-based pseudo viruses bearing the S protein were generated in Lenti-X™ 293 T cells (Takara Bio Inc.), and viral suspensions having the same viral RNA copies/mL were prepared. Two-fold serial dilutions of heat-inactivated sera were mixed with an equal volume of viral suspension and incubated for 1 h at 37°C. The mixtures of sample and virus were added in duplicates to HEK293T, which stably expresses human ACE2 and TMPRSS2 cells, followed by incubating the plates at 37°C with 5% CO_2_ for 2 days. Cells were subjected to luciferase assay, and the intensity of luminescence was measured by a microplate reader. Percent neutralization was calculated as the difference between relative light units (RLUs) of the virus control and test sample wells: % Neutralization = 100% × [1 − (RLU of sample well ÷ mean RLU of virus control wells in each plate)]. Duplicate of %Neutralization values between plates were averaged and handled as one data point. The dilution factor for achieving 50% neutralization (50% neutralization titer; NT_50_) was calculated with XLfit® software (version 5.3.1.3) (IDBS). When the percentage neutralization was less than 50% at the first dilution, the NT_50_ was calculated as half of the first dilution factor, with a minimum value of 20 assigned for analysis.

### Measurement of strain specific anti-RBD IgG antibody titer

Strain-specific anti-RBD titers were measured using an in-house ELISA method, as described in a previous study [7]. The coated antigens for the WT, XBB.1.5, JN.1, and KP.3 were WT SARS-CoV-2 (COVID-19) S protein RBD, His Tag (MALS verified) (ACRO Biosystems, Newark, DE, USA, Cat# SPD-C52H3), XBB SARS-CoV-2 Spike RBD Protein, His Tag (XBB.1.5/Omicron) (MALS verified) (ACRO Biosystems, Cat# SPD-C5242), JN.1 SARS-CoV-2 Spike RBD Protein, His Tag (JN.1/Omicron) (MALS verified) (ACRO Biosystems, Cat# SPD-C5249), and KP3 SARS-CoV-2 Spike RBD Protein, His Tag (KP3/Omicron) (HPLC-verified)(Sino Biological Inc., Beijing, China, Cat# 40592-V08H157), respectively. Following antigen coating and plate washing, serum samples were applied using a serial dilution method. Goat Anti-Human IgG Fc (HRP) preadsorbed (Abcam, Cambridge, UK, Cat# ab98624) was used as the secondary antibody. The titers of each sample were determined using an endpoint method with multiple serum dilutions and were calculated by assigning the OD value (OD 450 nm - 570 nm) to a calibration curve established based on the dose–response of an anti–His Tag antibody (Human mAb to 6x Histag [RMH01], Abcam, Cat# ab219465) reacting with the His Tag.

### Statistical analysis

The differences in neutralizing activity and anti-RBD titers before and after vaccination were evaluated using a paired t-test. The correlation coefficient (r) and coefficient of determination (R^2^) between neutralizing activity and anti-RBD titers were calculated using Pearson’s correlation coefficient. Statistical significance was defined as *p* < 0.05 in a two-tailed test. The response rate (RR) following vaccination was defined as the percentage of samples exhibiting a ≥4-fold increase. All statistical analyses were performed using R version 4.1.1 and RStudio version 2021.09.0.

## Results

### Measured samples and demographic characteristics

Fifty-six participants were initially enrolled, and two were excluded due to infection between the two sampling points. The demographic characteristics of the remaining 54 are shown in Table 1. The median age was 70.5 years, 15 (27.8%) were male, 8 (14.8%) had a history of past infection, and 13 (24.1%) had an unknown infection status. The majority of participants (41 of 54, 75.9%) had received more than four times of SARS-CoV-2 vaccinations, while the remaining 13 (24.1%) had an unknown vaccination status. The mean sampling day before vaccination was 4.9 days prior (95% confidence interval [CI]: −8.5 to −1.2 days), and the mean sampling day after vaccination was 33.9 days post-vaccination (95% CI: 30.9 to 36.9 days).

**Table 1.** Demographic Characteristics.

|  |  |  |
| --- | --- | --- |
| Total (n) |  | 54 |
| Sex |  |  |
|  | Male, n (%) | 15 (27.8) |
|  | Female, n (%) | 39 (72.2) |
| Age |  |  |
|  | Median (IQR), y | 70.5 (12.5) |
|  | Mean (95%CI), y | 67.9 (64.3 – 71.6) |
| Past infection |  |  |
|  | Yes (n (%)) | 8 (14.8) |
|  | No (n (%)) | 33 (61.1) |
|  | Unknown (n (%)) | 13 (24.1) |
| Vaccination history |  |  |
|  | 4 times (n (%)) | 3 (5.6) |
|  | 5 times (n (%)) | 7 (13.0) |
|  | 6 times (n (%)) | 31 (57.4) |
|  | Unknown (n (%)) | 13 (24.1) |
| Serum sampling point before vaccination |  |  |
|  | Median (IQR), d | 0 (0) |
|  | Mean (95%CI), d | -4.9 (-8.5 – -1.2) |
| Serum sampling point after vaccination |  |  |
|  | Median (IQR), d | 31.5 (12.8) |
|  | Mean (95%CI), d | 33.9 (30.9 – 36.9) |

### Correlation between neutralizing activity and anti-RBD IgG antibody titer

A scatter plot of neutralizing activity and anti-RBD titers is shown in Fig. 1. Among all samples, including pre- and post-vaccination samples, anti-RBD titers against the WT, XBB.1.5, JN.1, and KP.3 strains exhibited strong correlations with neutralizing activity (r = 0.94, 0.87, 0.86, and 0.82, respectively). When analyzing samples separately before and after vaccination, the correlation coefficient for all four strains remained high (r = 0.92, 0.83, 0.85, and 0.82, respectively, for pre-vaccination samples, and r = 0.96, 0.85, 0.82, and 0.75, respectively, for post-vaccination samples). The correlation remained for all four strains, both with and without a history of past infection (Fig. S1).

**Fig. 1.**
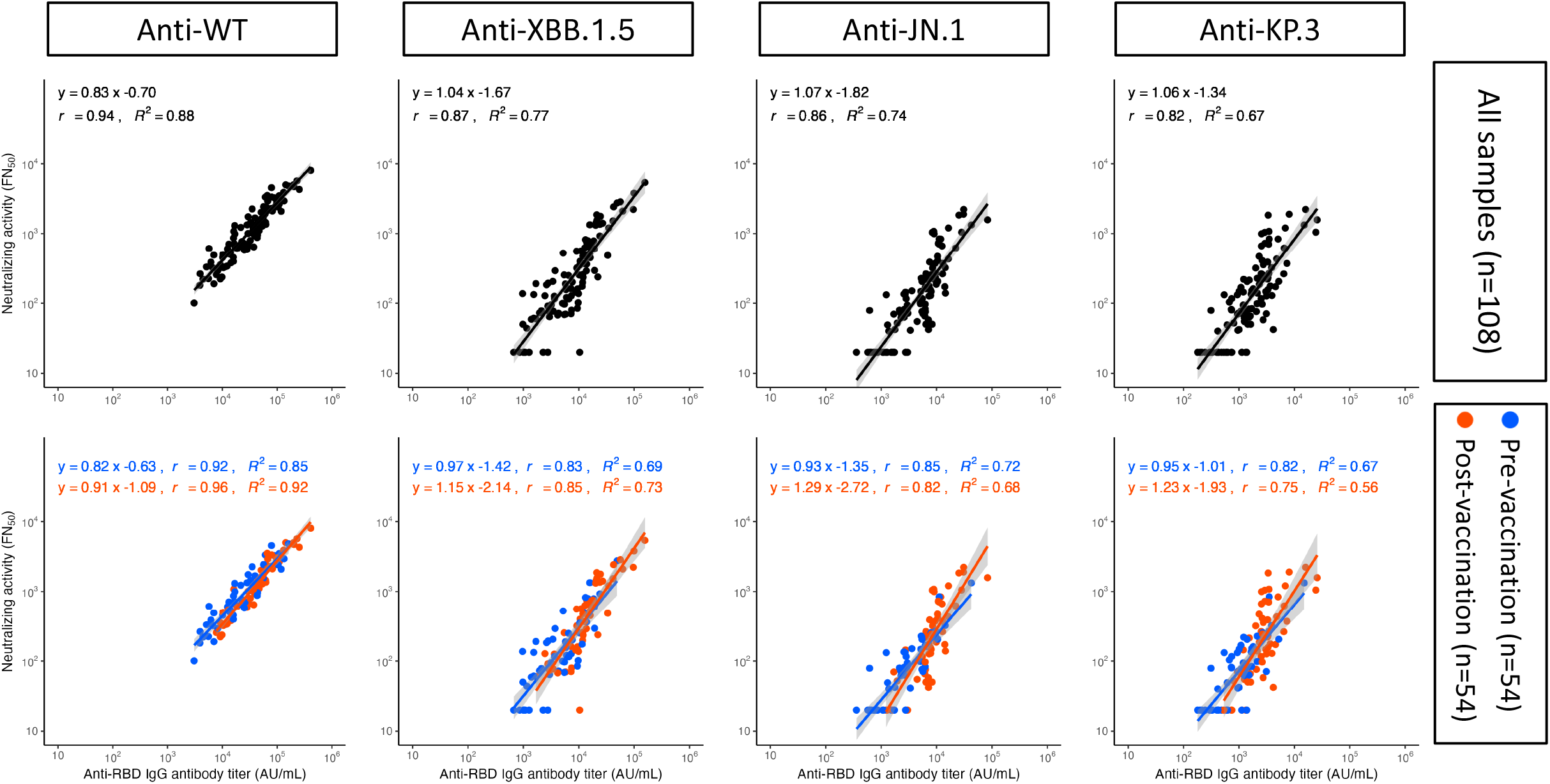
Correlation between neutralizing activity and anti-RBD IgG titers. The scatter plots illustrate the relation between neutralizing activity (50% neutralization titer; NT_50_) and anti-RBD IgG antibody titers (AU/mL) for the wild-type (WT), XBB.1.5, JN.1, and KP.3 strains. The black-filled circles in the upper part of the figure represent individual values. The blue-filled circles in the lower part of the figure represent pre-vaccination sample values, while the red-filled circles represent the post-vaccination sample values measured for each strain. The lines in each graph in the upper part of the figure represent regression lines, with the gray-shaded areas around them indicating the 95% confidence interval. In the lower panel of the figure, the blue and red lines represent regression lines for pre- and post-vaccination samples, respectively, with the gray-shaded areas indicating their 95% confidence interval. The equations displayed in the upper left corner of each section represent the regression equation, r (the correlation coefficient), and R^2^ (the coefficient of determination).

### Neutralizing activity before and after vaccination

A box plot of the NT_50_ values before and after XBB.1.5-adapted vaccination is shown in the upper part of Fig. 2. The geometric mean (GM) NT_50_ against the WT strain was 824 for pre-vaccination samples and 1,353 for post-vaccination samples, representing a 1.64-fold increase with an RR of 5.6%. The GM NT_50_ against the XBB.1.5 variant was 127 for pre-vaccination samples and 428 for post-vaccination samples, representing a 3.38-fold increase (RR = 35.2%). Similarly, the fold increase and RR values were 3.08-fold (27.8%) for the JN.1 variant and 2.44-fold (22.2%) for the KP.3 variant. The NT_50_ values were significantly increased in the post-vaccination samples of all four tested strains (*p* < 0.001).

**Fig. 2.**
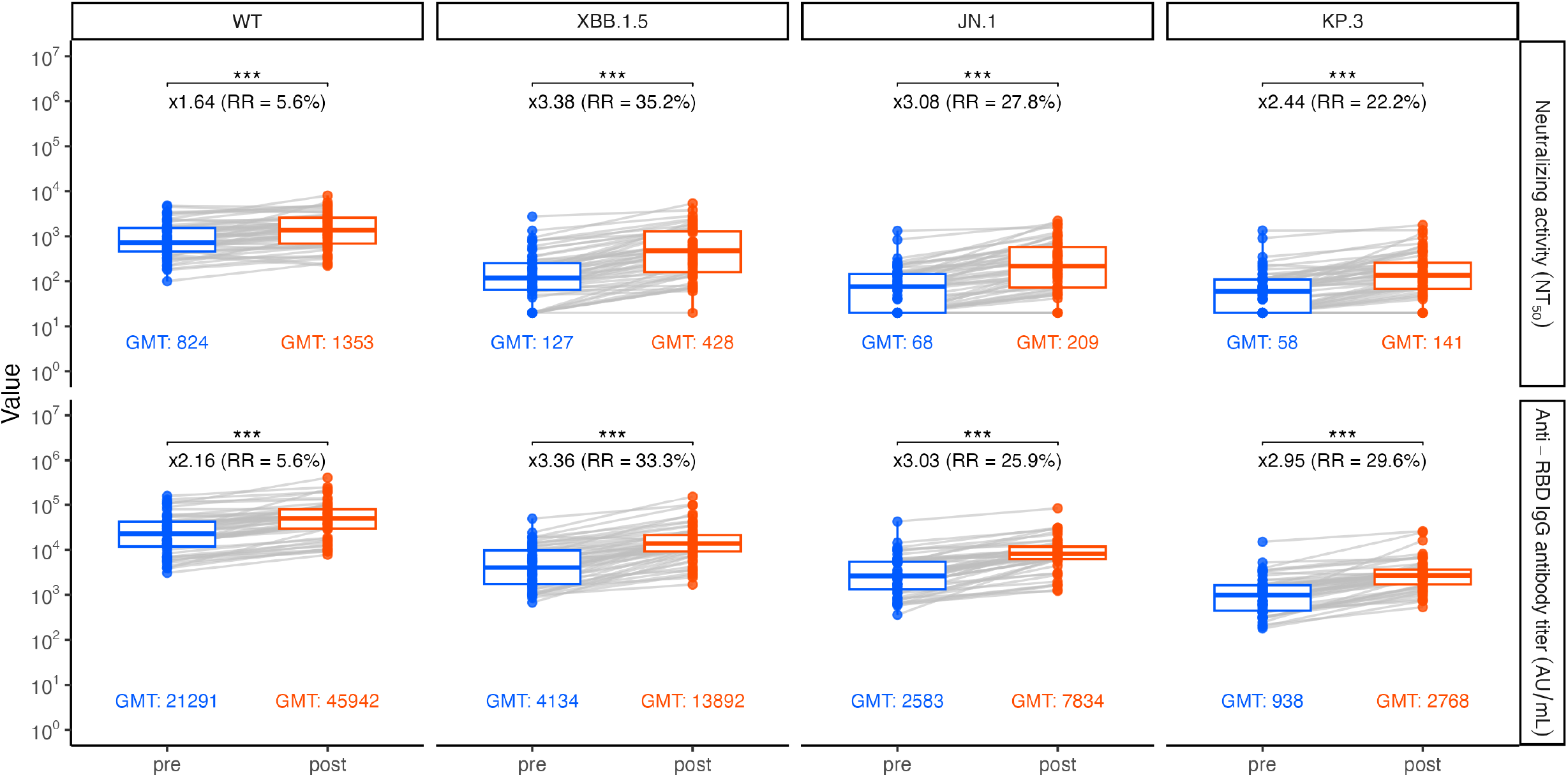
Neutralizing activity and anti-RBD antibody IgG titers before and after XBB.1.5-adapted vaccination. The Box-and-whisker plots depict neutralizing activity (50% neutralization titer; NT_50_) (upper part) and anti-RBD IgG antibody titers (AU/mL) (lower part) for the wild-type (WT), XBB.1.5, JN.1, and KP.3 strains. The blue plots represent individual values of pre-vaccination samples, while the red plots represent those of post-vaccination samples. The bottom and top of each box represent the first (Q1) and third quartiles (Q3), respectively, while the band near the middle of the box indicates the median. The ends of the whiskers represent the lowest and highest data points still within 1.5 times the interquartile range (IQR) of Q1 and Q3. Gray lines connect the pre- and post-vaccination data points of the same participant. The numbers below each box-and-whisker plot indicate the geometric mean (GM) value. Numbers marked with × above the box- and-whisker plots represent the fold change in GM value following vaccination compared to the pre-vaccination samples. RR, shown next to the fold change, represents the response ratio, defined as the percentage of samples exhibiting a ≥4-fold increase. *** denotes a paired t-test with p < 0.001.

### Anti-RBD IgG antibody titer before and after vaccination

A box plot of the anti-RBD IgG titers before and after XBB.1.5-adapted vaccination is shown in the lower part of Fig. 2. The geometric mean titer (GMT) against the WT strain was 21,291 for pre-vaccination samples and 45,942 for post-vaccination samples, representing a 2.16-fold increase (RR = 5.6%). The GMT against the XBB.1.5 variant was 4,134 for pre-vaccination samples and 13,892 for post-vaccination samples, representing a 3.36-fold increase (RR = 33.3%). Similarly, the GMT fold increase and RR values were 3.03-fold (25.9%) for the JN.1 variant and 2.95-fold (29.6%) for the KP.3 variant. The GMT values were significantly increased in the post-vaccination samples of all four tested strains (*p* < 0.001).

### Ratio of variants to WT before and after vaccination

To assess variant specificity, a box plot was done of the ratios of the XBB.1.5, JN.1, and KP.3 variants to the WT strain for neutralizing activity and anti-RBD titers, as shown in Fig. 3. For neutralizing activity, the mean ratio of the XBB.1.5 variant to the WT strain before vaccination was 0.19 and increased significantly to 0.36 after vaccination (1.92-fold, *p* < 0.001). Similarly, the fold increases in the ratios of the JN.1 and KP.3 variants to the WT strain were 1.95-fold and 1.48-fold, respectively (*p* < 0.001). A significant increase in anti-RBD titers was also observed for all three variants, with fold changes in the ratios of the XBB.1.5, JN.1, and KP.3 variants to the WT strain of 1.47-fold, 1.41-fold, and 1.38-fold, respectively (*p* < 0.001).

**Fig. 3.**
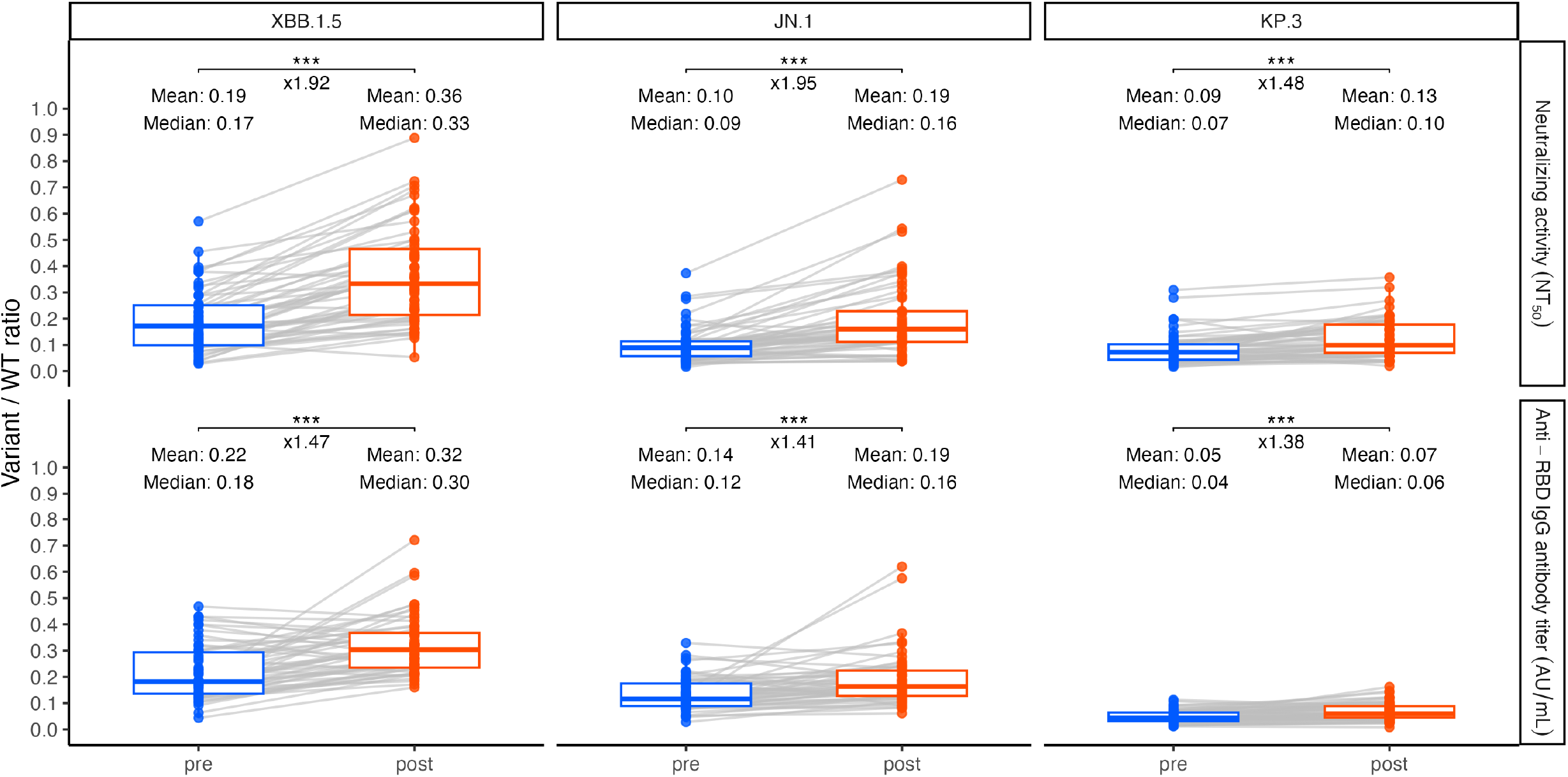
Ratio of variant values to wild-type before and after XBB.1.5-adapted vaccination. The box-and-whisker plots depict the ratio of the XBB.1.5, JN.1, and KP.3 variants to the wild-type (WT) strain for neutralizing activity (50% neutralization titer; NT_50_) (upper panel) and anti-RBD IgG antibody titers (AU/mL) (lower panel). The blue plots represent individual values of pre-vaccination samples, while the red plots represent those of post-vaccination samples. The bottom and top of each box represent the first (Q1) and third quartiles (Q3), respectively, while the band near the middle of the box indicates the median. The ends of the whiskers represent the lowest and highest data points still within 1.5 times the interquartile range (IQR) of Q1 and Q3. Gray lines connect the pre- and post-vaccination data points from the same participant. The numbers below each box-and-whisker plot indicate the geometric mean (GM) value. Numbers above the box-and-whisker plots represent the mean and median of the ratio, while numbers marked with × indicate the fold change in the mean value following vaccination compared to pre-vaccination samples. *** denotes a paired t-test with p < 0.001.

## Discussion

The RBD has been identified as critical region of SARS-CoV-2 that facilitates viral entry into host cells via ACE-2 receptor [17,18]. By interfering with this function, RBD-specific antibodies are reported to mediate approximately 90% of the neutralizing activity in convalescent sera [19]. Previous studies reported a strong correlation between neutralizing activity and anti-RBD titers against the WT strain following WT-targeted vaccination [20,21]. On the other hand, some studies have reported that anti-RBD titers measured using the WT strain show a substantially reduced correlation with neutralizing activity against the Omicron variant [22,23]. Considering this issue, we measured strain-specific anti-RBD antibodies in this study to provide a more accurate assessment.

The correlation between neutralizing activity and anti-RBD titers against WT remained significantly high (r > 0.9) in both the pre- and post-vaccination samples of this study (Fig. 1). This result suggests that exposure to a novel variant, such as XBB.1.5, does not create a discrepancy between neutralizing activity and anti-RBD titers against WT substantial enough to disrupt their correlation. Strong correlations were also observed for the XBB.1.5, JN.1, and KP.3 variants in both pre- and post-vaccination samples. These findings imply that anti-RBD IgG binding antibodies continue to play an important role in neutralization, not only against the WT strain but also against the XBB.1.5, JN.1, and KP.3 variants, despite the presence of significant mutations in the RBD. Additionally, past infection history did not affect the correlation (Fig. S1).

The WT strain exhibited the highest neutralizing activity and anti-RBD IgG titer in pre-vaccination samples, with further increases observed in post-vaccination samples (Fig.2). This result suggests that antibodies with neutralizing activity against the WT strain remained dominant in these individuals and that they are stimulated following XBB.1.5-adapted vaccination. Although the previous bivalent vaccines targeting BA.1 or BA.4/5 also included mRNA for the WT strain, the XBB.1.5-adapted vaccine was the first mRNA vaccine administered in Japan that did not contain mRNA for the WT strain. Nevertheless, the stimulation of WT-specific antibodies following XBB.1.5-adapted vaccination may be attributed to a mechanism similar to that observed after BA.1 infection through which antibodies targeting epitopes shared by the pre-immunized WT strain and newly exposed variants play a significant role [24,25]. On the other hand, the fold change and RR values in neutralizing activity and anti-RBD titers from pre-to post-vaccination was highest against the XBB.1.5 strain, potentially reflecting the generation of de novo immunity that is independent of prior exposure to earlier strains but that is elicited following XBB.1.5-adapted vaccination. The significant increases observed in the JN.1 and KP.3 variants may indicate that strain-targeted mRNA vaccination can elicit binding antibodies with neutralizing activity that are effective not only against the targeted strain but also, to some extent, against genetically distinct lineages. The analysis of variant specificity, represented as the ratio to the WT strain (Fig. 3), showed a significant increase for the XBB.1.5 variant following XBB.1.5-adapted vaccination. This increase was also observed for the JN.1 and KP.3 variants; however, the fold change tended to diminish for the KP.3 variant. This trend may be due to antigenic differences between KP.3 and XBB.1.5, as documented in a previous study [26].

This study has several limitations. First, the sample was not standardized. All participants were Japanese, only 27.8% were male, and the number of children and young adults included was very small. Second, we were unable to stratify the participants based on their past infection and vaccination histories due to the small sample size. These factors may have influenced the results for neutralizing activity and anti-RBD titers.

In summary, we found that strain-specific anti-RBD IgG antibody titers could serve as a less labor-intensive and more cost-effective indicator of humoral immunity, demonstrating a strong correlation with neutralizing activity not only for the WT strain but also for the XBB.1.5, JN.1, and KP.3 variants in individuals vaccinated with the XBB.1.5-adapted vaccine. Background of population concerning COVID-19 vaccination and infection is becoming more diverse. The measurement of strain-specific anti-RBD IgG antibody titers in a large number of samples is expected to provide valuable data not only on differences in antibody responses induced by newly adapted vaccines among variants but also on the impact of patient background factors such as past infections. The obtained information may contribute to the development of future vaccines.

## Data Availability

All data produced in the present study are available upon reasonable request to the authors

## Author Contributions

TG, TS, NT, YC, and HI contributed to the study design and conducted the investigation. NK and TB were responsible for sample collection and data extraction from patient charts. TG and TS developed the protocol and conducted the measurement of anti-RBD titers using an in-house ELISA. SS and SI performed the measurement of neutralizing activity using a pseudovirus assay. YC and HI supervised the methodology and analyses. TG drafted the manuscript and had final responsibility for the decision to submit it for publication. All authors critically reviewed and approved the final manuscript.

## Acknowledgments

We thank the following doctors for participating in this study: Dr. Kenichi Doniwa, Dr. Mariko Echizen, Dr. Woon Joo Lee.

---

**Fig. S1.**
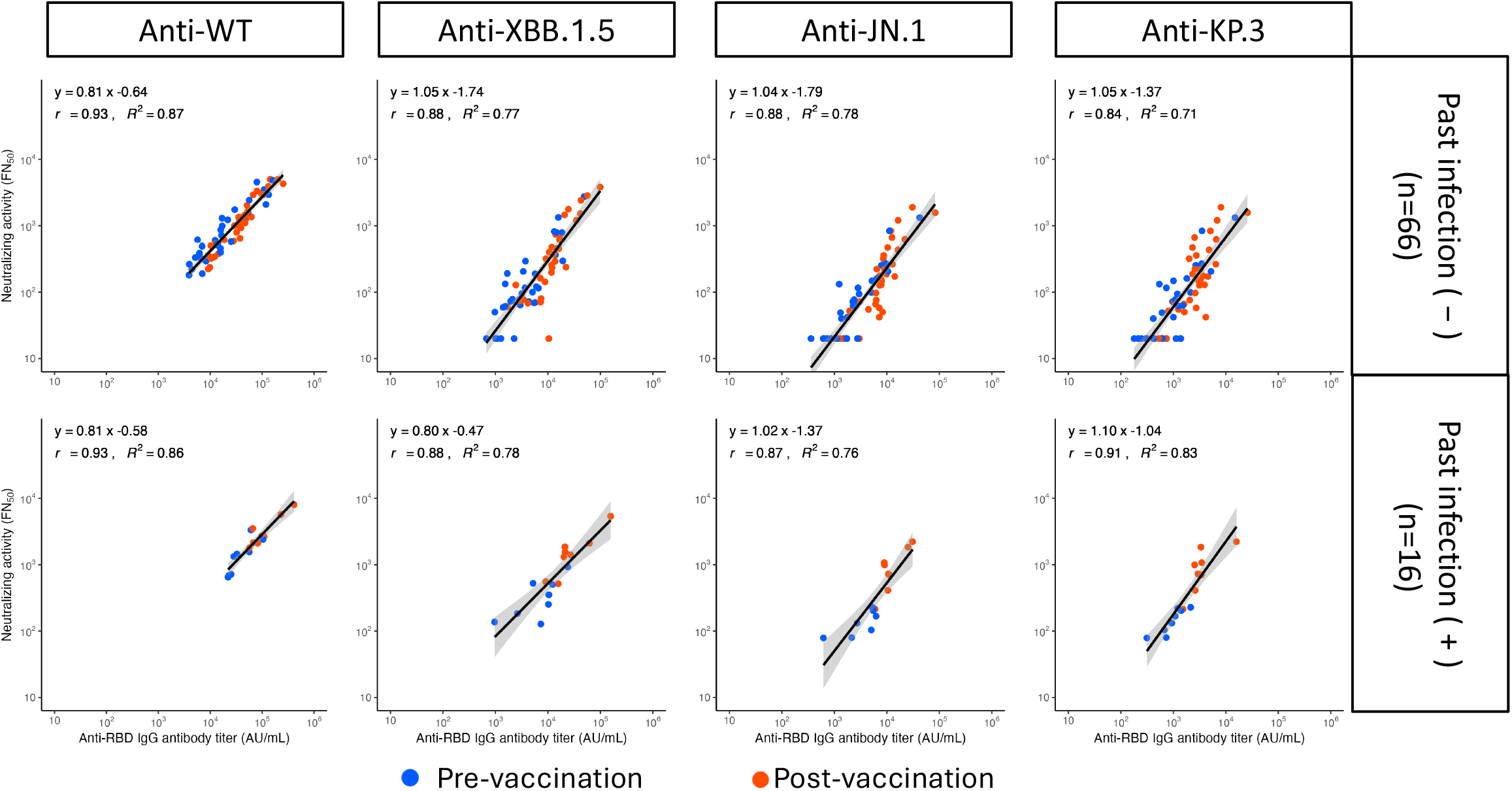
Correlation between neutralizing activity and anti-RBD IgG titers stratified by past infection history. The scatter plots illustrate the relation between the neutralizing activity (50% neutralization titer; NT_50_) and anti-RBD IgG antibody titers (AU/mL) of individuals without a history of past infection (upper panel) and those with (lower panel). Each plot in the figure represents individual values for the wild-type (WT), XBB.1.5, JN.1, and KP.3 strains. The blue-filled plots indicate pre-vaccination sample values, while the red-filled plots indicate the post-vaccination sample values measured for each strain. The lines in each graph represent regression lines, with the gray-shaded areas around them indicating the 95% confidence intervals. The equations displayed in the upper left corner of each section represent the regression equation, r (the correlation coefficient), and R^2^ (the coefficient of determination).

## References

[1] Ju B, Zhang Q, Ge J, Wang R, Sun J, Ge X, et al. Human neutralizing antibodies elicited by SARS-CoV-2 infection. Nature 2020;584:115–9.

[2] Garcia-Beltran WF, Lam EC, Astudillo MG, Yang D, Miller TE, Feldman J, et al. COVID-19-neutralizing antibodies predict disease severity and survival. Cell 2021;184:476-488.e11.

[3] Morinaga Y, Tani H, Terasaki Y, Nomura S, Kawasuji H, Shimada T, et al. Correlation of the commercial anti-SARS-CoV-2 receptor binding domain antibody test with the chemiluminescent reduction neutralizing test and possible detection of antibodies to emerging variants. Microbiol Spectr 2021;9:e0056021.

[4] Salazar E, Kuchipudi SV, Christensen PA, Eagar T, Yi X, Zhao P, et al. Convalescent plasma anti-SARS-CoV-2 spike protein ectodomain and receptor-binding domain IgG correlate with virus neutralization. J Clin Invest 2020;130:6728–38.

[5] Decru B, Van Elslande J, Steels S, Van Pottelbergh G, Godderis L, Van Holm B, et al. IgG anti-spike antibodies and surrogate neutralizing antibody levels decline faster 3 to 10 months after BNT162b2 vaccination than after SARS-CoV-2 infection in healthcare workers. Front Immunol 2022;13:909910.

[6] Higashimoto Y, Kozawa K, Miura H, Kawamura Y, Ihira M, Hiramatsu H, et al. Correlation between anti-S IgG and neutralizing antibody titers against three live SARS-CoV-2 variants in BNT162b2 vaccine recipients. Hum Vaccin Immunother 2022;18:2105611.

[7] Goto T, Chong Y, Tani N, Susai N, Yoshinaga T, Sasaki T, et al. Distinct features of SARS-CoV-2 humoral immunity against Omicron breakthrough infection. Vaccine 2023;41:7019–25.

[8] Nextstrain_Japan. Nextstraion n.d. https://nextstrain.org/ncov/open/asia/all-time?c=country&f_country=Japan&lang=ja&p=grid&r=division&tl=pango_lineage&transmissions=show (accessed February 11, 2025).

[9] Qu P, Faraone JN, Evans JP, Zheng Y-M, Carlin C, Anghelina M, et al. Enhanced evasion of neutralizing antibody response by Omicron XBB.1.5, CH.1.1, and CA.3.1 variants. Cell Rep 2023;42:112443.

[10] Uriu K, Ito J, Zahradnik J, Fujita S, Kosugi Y, Schreiber G, et al. Enhanced transmissibility, infectivity, and immune resistance of the SARS-CoV-2 omicron XBB.1.5 variant. Lancet Infect Dis 2023;23:280–1.

[11] Kaku Y, Okumura K, Padilla-Blanco M, Kosugi Y, Uriu K, Hinay AA Jr, et al. Virological characteristics of the SARS-CoV-2 JN.1 variant. Lancet Infect Dis 2024;24:e82.

[12] Lewnard JA, Mahale P, Malden D, Hong V, Ackerson BK, Lewin BJ, et al. Immune escape and attenuated severity associated with the SARS-CoV-2 BA.2.86/JN.1 lineage. Nat Commun 2024;15:8550.

[13] Li P, Faraone JN, Hsu CC, Chamblee M, Liu Y, Zheng Y-M, et al. Neutralization and stability of JN.1-derived LB.1, KP.2.3, KP.3 and KP.3.1.1 subvariants. BioRxivorg 2024:2024.09.04.611219. 10.1101/2024.09.04.611219.

[14] Springer DN, Camp JV, Aberle SW, Deutsch J, Lammel O, Weseslindtner L, et al. Neutralization of SARS-CoV-2 Omicron XBB.1.5 and JN.1 variants after COVID-19 booster-vaccination and infection. J Med Virol 2024;96:e29801.

[15] Wang Q, Guo Y, Bowen A, Mellis IA, Valdez R, Gherasim C, et al. XBB.1.5 monovalent mRNA vaccine booster elicits robust neutralizing antibodies against XBB subvariants and JN.1. Cell Host Microbe 2024;32:315-321.e3.

[16] Fujitani M, Lu X, Shinnakasu R, Inoue T, Kidani Y, Seki NM, et al. Longitudinal analysis of immune responses to SARS-CoV-2 recombinant vaccine S-268019-b in phase 1/2 prime-boost study. Front Immunol 2025;16:1550279.

[17] Wrapp D, Wang N, Corbett KS, Goldsmith JA, Hsieh C-L, Abiona O, et al. Cryo-EM structure of the 2019-nCoV spike in the prefusion conformation. Science 2020;367:1260–3.

[18] Shang J, Ye G, Shi K, Wan Y, Luo C, Aihara H, et al. Structural basis of receptor recognition by SARS-CoV-2. Nature 2020;581:221–4.

[19] Lapuente D, Winkler TH, Tenbusch M. B-cell and antibody responses to SARS-CoV-2: infection, vaccination, and hybrid immunity. Cell Mol Immunol 2024;21:144–58.

[20] Younes S, Nicolai E, Pieri M, Bernardini S, Daas HI, Al-Sadeq DW, et al. Follow-up and comparative assessment of SARS-CoV-2 IgA, IgG, neutralizing, and total antibody responses after BNT162b2 or mRNA-1273 heterologous booster vaccination. Influenza Other Respi Viruses 2024;18:e13290.

[21] Güzel I, Öztürk G, Appak Ö, Çağlayan D, Süner AF, Irmak Ç, et al. Neutralizing and binding antibody dynamics following primary and booster COVID-19 vaccination among healthcare workers. BMC Infect Dis 2025;25:218.

[22] Springer DN, Perkmann T, Jani CM, Mucher P, Prüger K, Marculescu R, et al. Reduced sensitivity of commercial Spike-specific antibody assays after primary infection with the SARS-CoV-2 Omicron variant. Microbiol Spectr 2022;10:e0212922.

[23] Dépéry L, Bally I, Amen A, Némoz B, Buisson M, Grossi L, et al. Anti-SARS-CoV-2 serology based on ancestral RBD antigens does not correlate with the presence of neutralizing antibodies against Omicron variants. Microbiol Spectr 2025;13:e0156824.

[24] Quandt J, Muik A, Salisch N, Lui BG, Lutz S, Krüger K, et al. Omicron BA.1 breakthrough infection drives cross-variant neutralization and memory B cell formation against conserved epitopes. Sci Immunol 2022;7:eabq2427.

[25] Kaku CI, Bergeron AJ, Ahlm C, Normark J, Sakharkar M, Forsell MNE, et al. Recall of preexisting cross-reactive B cell memory after Omicron BA.1 breakthrough infection. Sci Immunol 2022;7:eabq3511.

[26] Roederer AL, Cao Y, St Denis K, Sheehan ML, Li CJ, Lam EC, et al. Ongoing evolution of SARS-CoV-2 drives escape from mRNA vaccine-induced humoral immunity. Cell Rep Med 2024;5:101850.

